# Obesity as a predictor for adverse outcomes among COVID-19 patients: A meta-analysis

**DOI:** 10.1101/2020.11.27.20239616

**Authors:** Pranta Das, Nandeeta Samad, Abdul-Aziz Seidu, Richard Gyan Aboagye, Justice Kanor Tetteh, Bright Opoku Ahinkorah

**Author notes:** Email Addresses PD.

## Abstract

**Background:** This meta-analysis sought to determine the estimated association between obesity and adverse outcomes among COVID-19 patients.

**Methods:** We followed the recommended PRISMA guidelines. A systematic literature search was conducted in PubMed, Google Scholar, and ScienceDirect for published literature between December 1, 2019, and October 2, 2020. The data for the study were pooled from studies that contained the search terms “Obesity” AND (COVID-19 or 2019-nCoV or Coronavirus or SARS-CoV-2) AND (“ICU admission” OR “Hospitalization” OR “Disease severity” OR “Invasive mechanical ventilator” OR “Death” OR “Mortality”). All the online searches were supplemented by reference screening of retrieved studies for additional literature. The pooled odds ratio (OR) and confidence intervals (CI) from the retrieved studies were calculated using the random effect model (Inverse-Variance method).

**Findings:** Five studies with a combined sample size of 335,192 patients were included in the meta-analysis. The pooled OR from the final analysis showed that patients who are severely obese were more likely to experience adverse outcome (death or ICU admission or needing IMV or hospitalization) compared to the normal patients [OR = 2.81, 95% CI = 2.33 – 3.40, I^2^ = 29%].

**Conclusion:** Severe obesity is a risk factor in developing adverse outcomes among COVID-19 patients. The finding of the study signifies promotive, preventive, and curative attention to be accorded patients diagnosed with severe obesity and COVID-19.

## Background

The World Health Organization (WHO) declared the severe acute respiratory syndrome coronavirus diseases 2019 (COVID-19) a public health emergency of international concern (PHEIC) on the 30^th^ of January 2020 and further characterized the spread of the novel COVID-19 disease a global pandemic on the 11^th^ of March, 2020 [1]. As at 30^th^ January 2020, there were 7818 total confirmed cases globally with only 82 cases recorded outside China – the epicenter of the pandemic [2]. Between 30^th^ January 2020 and 20^th^ October 2020, over 40.3 million confirmed cases across 217 countries with over 1.12 million deaths was reported globally making the pandemic one of the deadliest in the past two centuries [3].

Initial findings from early studies pointed towards age with older individuals being particularly at risk, those with non-communicable diseases (NCDs), such as, diabetes mellitus or cardiovascular diseases such as hypertension, respiratory conditions, and or kidney disease [4]. Further studies have led to the discovery that obesity is a plausible risk factor for severe illness, hospitalization and even death from COVID-19 [5, 6].

Obesity is known to be a risk factor for many NCDs [7] and other medical conditions leading to an increased risk of hospitalization, serious illness, and mortality [8]. In 2009, for example, obesity was highlighted as an independent risk factor for elevated disease severity and mortality for the HINI influenza virus [8-9] and even though the association between obesity and increased disease severity is not novel, empirical knowledge about its association with the novel COVID-19 virus pandemic is important to guide the fast changing efforts being adopted by health systems globally to halt its spread, severe illness, hospitalizations and mortalities. Increasingly, obesity is being identified as a predisposing factor for COVID-19 and this is particularly worrying as obesity affects a large proportion of the world’s population as 39% of adults globally are overweight (BMI ≥25.0 to 29.9 kg/m^2^) and 13% have clinical obesity (BMI ≥30.0 kg/m^2^) [11, 12].

The prevalence of persons with obesity globally is at an all-time high for both higher income countries and low-and middle-income countries [13]. According to Jones-Smith et al [14] and Jones-Smith et al [15], more than 70% of persons with overweight/obesity live in low or middle□income countries, and as a country’s economy grows, the burden of persons with obesity shifts to the poor. More worrying is the fact that empirical evidence shows that obesity leads to impaired immunity, chronic inflammation, decreasing lung capacity and reserve making ventilation more difficult, blood that’s prone to clot, and lower vaccine response as recorded previously for some diseases such as influenza, hepatitis B and tetanus, and all of which can worsen COVID-19 illness [12, 16-18]. Moreover, obesity and NCDs in the geriatric population are significantly associated with polypharmacy, which suppresses immunity leading to increased risk of morbidity and mortality in acute infections that occur in COVID-19 patients [19]. These makes obesity especially in an era of a global pandemic, an important phenomenon worth examining.

There are growing empirical literature on the relationship between obesity and COVID-19. For example, in an observational cohort study in the UK which sampled more than 20,000 hospital in-patients with COVID-19, researchers found that obesity was an independent risk factor for high mortality on COVID-19 patients [20]. In another study in France, the prevalence of obesity among adult inpatients with COVID-19 was found to be high and obesity together with other comorbidities were significant in predicting adverse effect and even death for COVID-19 [21]. A study by Tartof, Qian and Hong [22] has also reported that obesity is a risky predisposing factor for persons who contract COVID-19, while adding that from their study findings obesity stood out from racial, ethnic, or socioeconomic factors when they were controlled for. In that same study, Tartof and colleagues found that data from the 6916 sampled patients in the study revealed that compared with those at normal body mass index (BMI) of 18.5 to 24 kg/m^2^, the risk of death more than doubled for COVID-19 patients with a body mass index (BMI) of 40 to 44 kg/m^2^ and nearly doubled again for those with a BMI of 45 kg/m^2^. This finding by Tartof and colleagues has been corroborated recently by a study in Saudi Arabia by Herbst et al. [23] who also found that obesity increases the risk of death from COVID-19 by 48%, the risk of hospitalization by 113%, and of needing intensive care by 74%. Herbst et al. [23] further highlighted a concern that a COVID-19 vaccine may not work for obese people due to the fact that flu vaccines don’t work effectively in people with BMI of more than 30.

Despite the growing evidence on the effect of obesity on COVID-19 treatment outcomes, it is important to note that in the past there have been some conflicting findings in relation to obesity and their effects on treatment outcomes for some respiratory diseases. For example, findings from a study in the United States which used almost 10,000 cases of seasonal influenza did not find any significant evidence of obesity as a risk factor for requiring mechanical ventilation or death [24] unlike the case in 2009 [25]. Furthermore, Umbrello et al. [26] have also reported that obesity was associated with increased survival rate for patients with acute respiratory distress syndrome. These conflicting reports in the past makes it relevant today for a strong synthesis and a meta-analysis of evidence to be established for obesity in the global fight against the COVID-19 pandemic.

Literature is littered with scattered pieces of evidence on how obesity put one at risk for adverse outcome of COVID-19. With this growing body of empirical literature owing to the rapid spread of the novel COVID-19 virus with its accompanying mortalities and the increasing growth in the proportion of the world’s population living with obesity, it has become imperative for this study to be conducted to synthetize the evidence, assess the strength of the evidence and obtain a single summary estimate of the effect of obesity on COVID-19 outcomes through a meta-analysis of empirical literature. This study is expected to provide conclusive empirical evidence to guide ongoing measures and programs put in place by health systems globally to halt the spread, hospitalizations and mortalities as a result of the COVID-19 pandemic.

## Methods

### Search strategy

A systematic literature search was conducted through databases PubMed, Google Scholar and ScienceDirect for English language literature published between December 1, 2019 and October 2,2020. The search consisting of the terms “Obesity” AND (COVID-19 or 2019-nCoV or Coronavirus or SARS-CoV-2) AND (“ICU admission” OR “Hospitalization” OR “Disease severity” OR “Invasive mechanical ventilator” OR “Death” OR “Mortality”) had been used to find potential papers. More details of the search in each database are presented in Table S1-S3 in the supplementary file.

### Eligibility criteria

The study which categorized BMI as normal (< 25 kg/m2), overweight (from 25 to < 30 kg/m2), moderate obesity (from 30 to < 35 kg/m2), and severe obesity (≥ 35 kg/m2) and at the same time reported outcomes such as ICU admission or requiring Invasive Mechanical Ventilation (IMV)or requiring hospitalization or death among the real-time reverse transcriptase-polymerase chain reaction (RT-PCR) or laboratory-confirmed COVID-19 patients were included. Only English language research articles published in a peer-reviewed journals were included in this study. Unpublished articles as well as preprints were excluded in the screening process. The studies with only cohort study design were included. Therefore, studies with other study designs, review articles, letters to the editor etc. were excluded. And the studies which reported odds ratio were only included. So, the studies reporting risk ratio (RR), hazard ratio (HR) etc. were excluded.

### Data extraction and study quality assessment

After selecting the relevant articles using the inclusion, exclusion criteria and following the PRISMA process, data were extracted from the selected articles. From each article extracted data included name of the first author, study design, country, follow-up period, sample size, age of the participants, outcome investigated and odds ratio associated with BMI category >=35 kg/m2, keeping BMI category <25 kg/m2 as the reference category, as well as the confidence interval associated with the odds ratio. Newcastle-Ottawa technique for the assessment of study quality for cohort study were used to assess the study quality of the included cohort studies. Studies with quality score 6 or more were included in the analysis.

### Statistical analysis

After extracting the necessary data pooled odds ratio (OR) and 95% confidence interval (CI) from the included studies were calculated using random effect model (Inverse-Variance method). Random effect model was selected regardless of the degree of heterogeneity among the included studies as the result from random effect model is generalizable. Between-study heterogeneity was assessed by calculating I^2^ statistic. Forrest plot was drawn to comprehensively present the information about included studies and between study heterogeneity. Egger’s bias test as well as funnel plot was drawn to assess the risk of publication bias. And finally, sensitivity analysis using sequential omission of included studies was performed to assess the consistency of the result. All the p-values less than 0.05 were considered significant. All the analysis was done using software STATA 12.

## Results

### Search results

A total of 2998 articles were identified through online database searching among which 340 were through PubMed, 2600 were through Google Scholar and 58 were through ScienceDirect (Figure 1). After screening title and abstract of those 2998 articles 18 were selected for full text screening.

**Figure 1:**
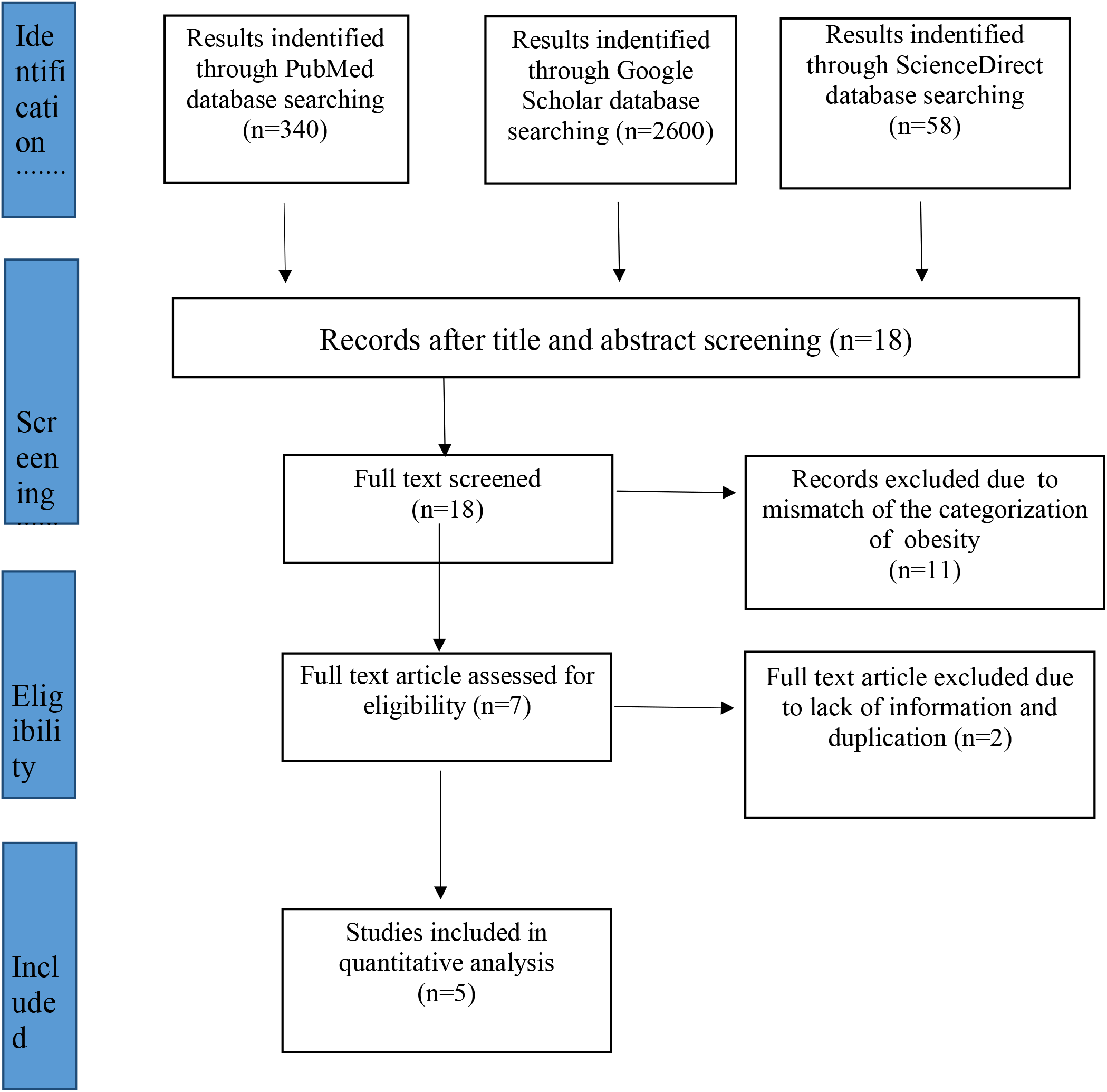
PRISMA flowchart for search strategy and the process of selecting articles

After the full text screening of those 18 articles, 11 articles were excluded due to mismatch of the categorization of the obesity. Then among the 7 articles, 2 articles were excluded due to lack of information and duplication. Finally, 5 articles were included for the quantitative analysis.

### Study Characteristics

Among the included studies, three studies were conducted in the USA, one in France and one in the UK. The study design of three studies among the five studies, were retrospective cohort, one was prospective cohort and one was population-based cohort study. The minimum sample size among the studies was 103 and maximum was 3,34,329. Two studies investigated the outcome ICU admission, one investigated needing Invasive Mechanical Ventilator (IMV), one investigated hospitalization, and one investigated death among the identified COVID-19 patients. All the included studies had good quality score (Table S4 in supplementary file). More characteristics of the included studies are presented in Table 1.

**Table 1:**
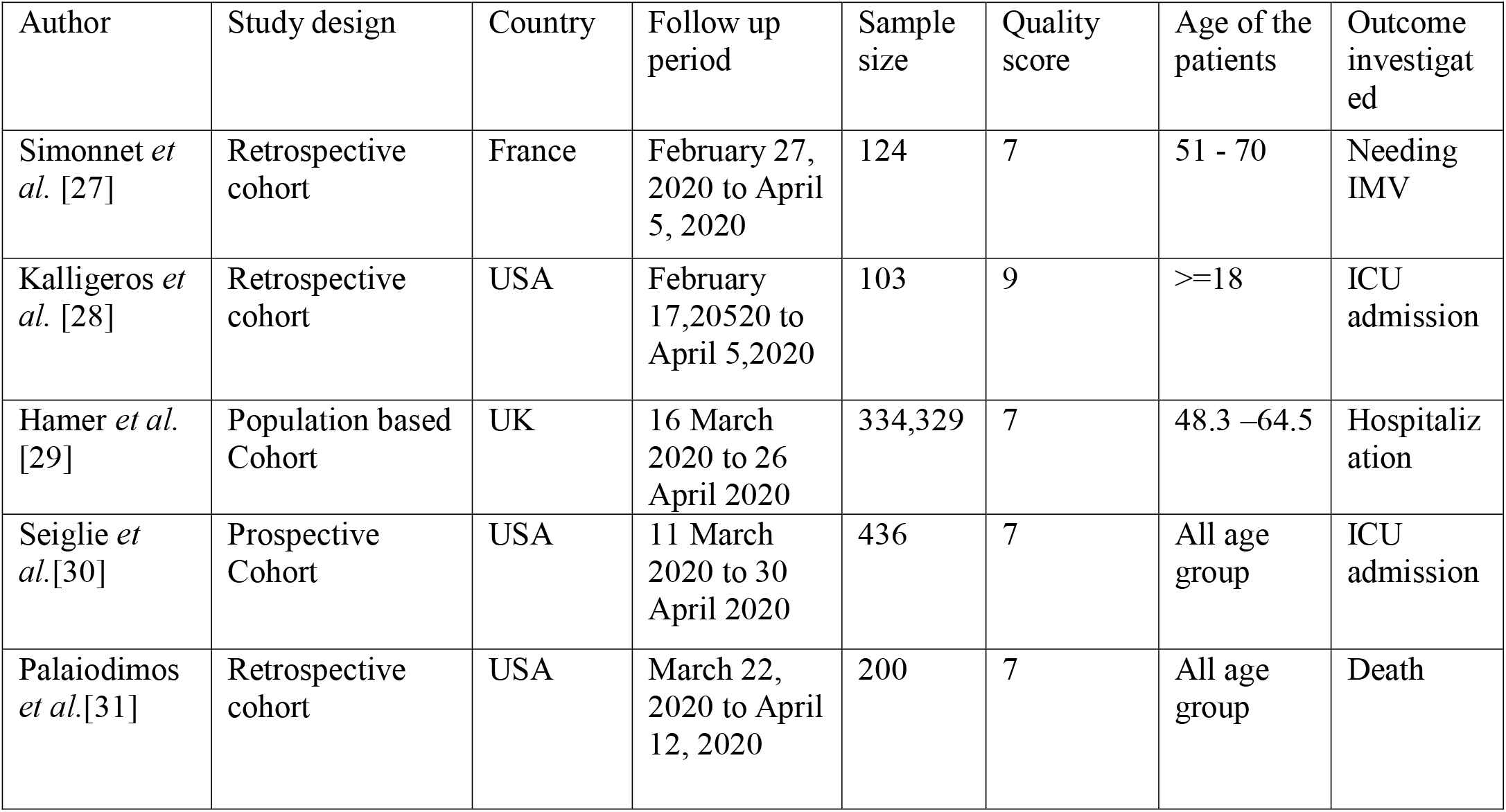
Characteristics of the study included in the analysis.

| Author | Study design | Country | Follow up period | Sample size | Quality score | Age of the patients | Outcome investigated |
| --- | --- | --- | --- | --- | --- | --- | --- |
| Simonnet *et al.* [27] | Retrospective cohort | France | February 27, 2020 to April 5, 2020 | 124 | 7 | 51 - 70 | Needing IMV |
| Kalligeros *et al.* [28] | Retrospective cohort | USA | February 17, 2020 to April 5, 2020 | 103 | 9 | >=18 | ICU admission |
| Hamer *et al.* [29] | Population based Cohort | UK | 16 March 2020 to 26 April 2020 | 334,329 | 7 | 48.3 –64.5 | Hospitalization |
| Seiglie *et al.* [30] | Prospective Cohort | USA | 11 March 2020 to 30 April 2020 | 436 | 7 | All age group | ICU admission |
| Palaiodimos *et al.* [31] | Retrospective cohort | USA | March 22, 2020 to April 12, 2020 | 200 | 7 | All age group | Death |

### Effect of severe obesity on adverse outcome in COVID-19 patients

The pooled OR for the adverse outcome among the COVID-19 patients, heterogeneity statistic and the result egger’s bias test of publication bias are presented in Table 2. The pooled OR obtained from the five studies suggesting that the people who are severely obese that is with BMI >=35 kg/m2 are 2.81 times more likely (OR=2.81, 95% CI= 2.33 – 3.40, I^2^=29%) to experience adverse outcome (death or ICU admission or needing IMV or hospitalization) compared to the normal people that is with BMI <25 kg/m2. The forest plot of the result is depicted in Figure 2. Publication bias was tested using egger’s bias test and creating funnel plot. The p-value associated with the egger’s bias test (p=0.873) suggests that no publication bias is present. Similar result can be seen through funnel plot. The funnel plot is presented in Figure S1 in the supplementary file.

**Table 2:** Summary effects of obesity on adverse outcome among COVID-19 infected persons and publication bias.

| Characteristic | Number of studies | Total number of patients | Summary statistics |  | Egger Bias test p-value |
| --- | --- | --- | --- | --- | --- |
|  |  |  | OR (95%CI) | Heterogeneity(I<sup>2</sup>) |  |
| Obesity | 5 | 335192 | 2.81(2.33 -3.40) | 29% | 0.873 |

Note: CI; Confidence interval

**Figure 2:**
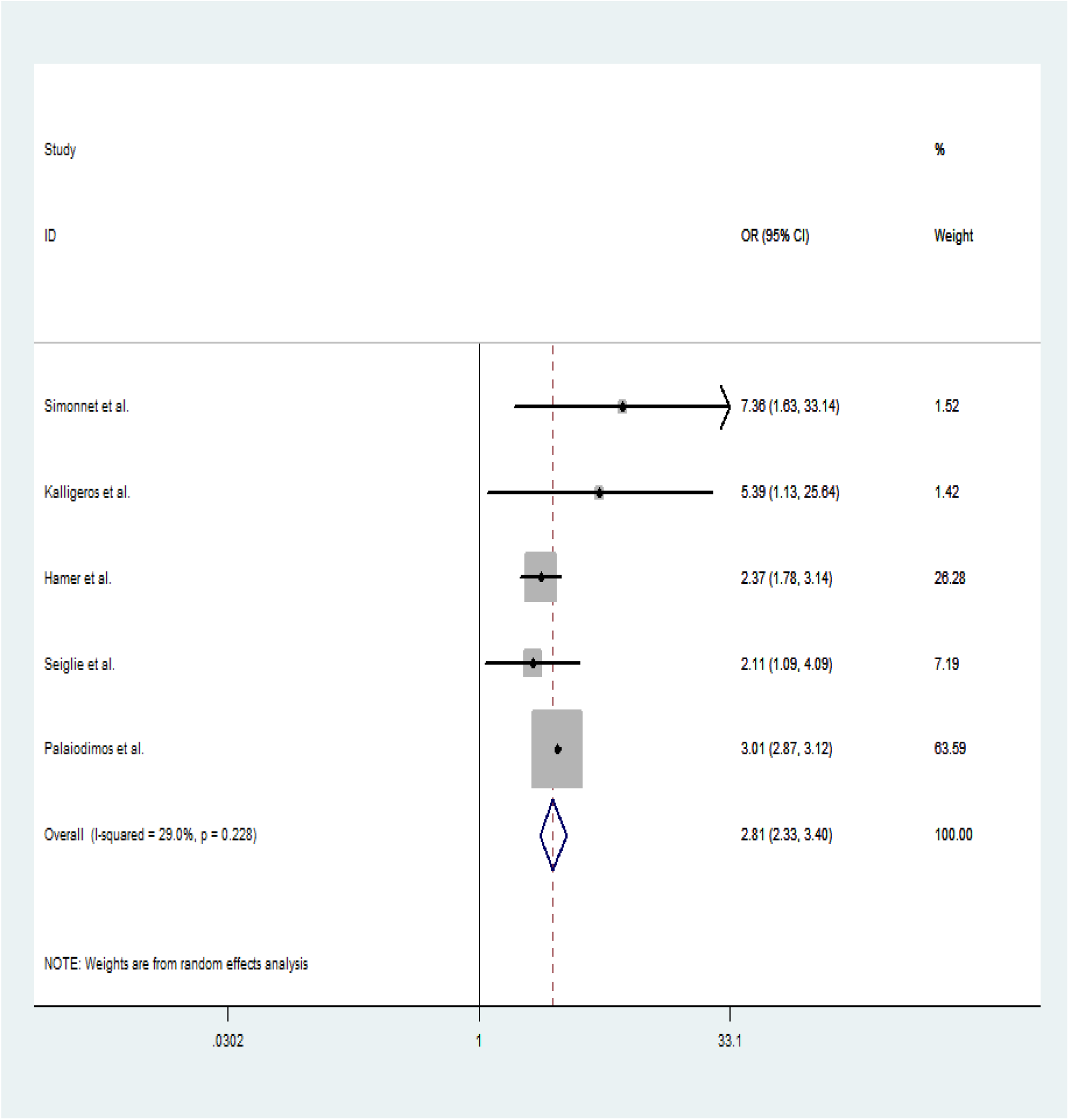
Forest plot of the result obtained using random effect model.

### Sensitivity analysis

The result of sensitivity analysis with the sequential omission of included studies is presented in Table 3. The result suggests that the pooled OR is not based on only one study; all the studies contributed to the final pooled result. The graph of the result obtained from sensitivity analysis is presented in figure S2 in the supplementary file.

**Table 3:** Result of sensitivity analysis.

| Study omitted | Estimate | 95% CI |  |
| --- | --- | --- | --- |
|  |  | Lower | Upper |
| Simonnet *et al.* | 2.7938795 | 2.34828 | 3.324034 |
| Kalligeros *et al.* | 2.7682662 | 2.2458508 | 3.4122028 |
| Hamer *et al.* | 3.0060809 | 2.8834586 | 3.1339176 |
| Seiglie *et al.* | 2.875937 | 2.3459527 | 3.5256522 |
| Palaiodimos *et al.* | 2.5062578 | 1.8378928 | 3.4176795 |
| Combined | 2.8145215 | 2.3324781 | 3.3961869 |

Note: CI; Confidence interval

## Discussion

The current study found that severely obese COVID-19 patients had higher odds of experiencing adverse outcomes such as death, ICU admission, needing IMV, and hospitalization. This study supports the evidence from clinical observations from [28, 32-35] that obesity is associated with increased risk of severe COVID-19, hospital admission, requiring IMV and death and intubation. The finding in our study confirms the association between obesity among patients and adverse effects (SARS-CoV-2 severity, requiring increased attention and intensive care for SARS-CoV-2) found in France [27].The result accords the earlier studies which showed that obesity is associated with the risk of increased hospitalisation, risk of developing adverse complications [36-39]. Similarly, obesity has been reported to be associated with a greater likelihood of enhancing transmission and developing adverse complications from disease such as influenza [40], influenza A (H1N1) infection [41-44].

Despite the significant findings shown in the present study, the exact mechanism necessitating the association between obesity and adverse effects of COVIDD-19 is not conclusive. However, several factors could account for the observed findings. First of all, the results could imply that patients with obesity have altered respiratory physiology, including decreased functional residual capacity, expiratory reserves volume, hypoxemia, and ventilation or perfusion abnormalities which could have predisposed them to contracting and developing severe complications from COVID-19 [28]. Secondly, the risk of obesity in patients with COVID-19 could have increased because the COVID-19 virus affects the adipose tissue which serves as a reservoir for human adenovirus Ad-36, influenza A virus, HIV, cytomegalovirus, Trypanosoma gondii, and Mycobacterium tuberculosis. This increases the tendency by which COVID-19 spread to other organs in the body, hence the several complications [45]. Also, the severe complications associated with COVID-19 among patients with obesity could result from the consequences of the underlying low-grade chronic inflammation, and suppression of innate and adaptive immune responses [33]. Another plausible explanation may be that mechanical dysfunction due to severe obesity could have increased the severity of lower respiratory tract infection and contribute to secondary infection [33].

### Strength and limitations of the study

This meta-analysis strongly reiterates the evidence that obesity is a risk factor for severe COVID-19 outcomes. However, the cross-sectional nature of the studies included in the present meta-analysis could limit the findings’ generalisability as causal inference is not drawn. Also, the studies included were from the USA, France, and the UK, which could limit its applicability due to differences in races, patterns of the severity of COVID-19 infections, and mortality rate.

## Conclusion

Our result showed that obesity is a contributory factor associated with increasing COVID-19 adverse outcomes. Among the patients with obesity, the risk of developing COVID-19 adverse outcomes was close to three folds. This finding underscores the need for obese people to strictly adhere to the COVID-19 prevention guidelines and minimal exposure to infected persons. Health promotion regarding physical exercise and balanced diet should be undertaken to prevent overweight and obesity. There is also the need for early detection and treatment of people with obesity diagnosed with COVID-19. Further research should focus on the factors influencing the association between obesity and the severity of COVID-19 outcomes.

## Data Availability

The data used to support the findings of this study are available from the corresponding author upon request.

## Abbreviations

BMI: Body Mass Index
COVID-19: Coronavirus Disease 2019
ICU: Intensive Care Unit
IMV: Invasive Mechanical Ventilator
HR: Hazard Ratio
PHEIC: Public Health Emergency of International Concern
NCDs: Non-Communicable Diseases
RR: Risk Ratio
SARS-COV-2: Severe Acute Respiratory Syndrome Coronavirus 2019.

## Authors’ contributions

NS and PD participated in conceptualization. Drafting the manuscript: NS, PD, AS, RGA, JKT and BOA. Revising the manuscript critically for important intellectual content: NS, PD, AS, RGA, JKT and BOA. All authors have read and approved the final manuscript.

## Acknowledgements

Not applicable.

## Consent for publication

Not applicable.

## Ethics approval and consent to participate

Not applicable

## References

1. WHO. WHO Director-General’s opening remarks at the media briefing on COVID-19 – 11 March 2020. 2020 Mar 11. Retrieved from www.who.int/dg/speeches/detail/who-director-general-s-opening-remarks-at-the-media-briefing-on-covid-19---11-march-2020.

2. WHO. Archived: WHO Timeline - COVID-19. 2020 Apr 27. Retrieved from: https://www.who.int/news/item/27-04-2020-who-timeline---covid-19 on 18/10/2020 at 1:21am

3. Worldometer. COVID-19 Coronavirus pandemic. Reported Cases and Deaths by Country, Territory, or Conveyance. 2020. Retrieved from: https://www.worldometers.info/coronavirus/?utm_campaign=homeAdUOA?Si on 18/10/2020 at 1:16am

4. Sattar N, McInnes IB, McMurray JJ. Obesity a risk factor for severe COVID-19 infection: multiple potential mechanisms. Circulation. 2020 Apr 22. https://doi.org/10.1161/CIRCULATIONAHA.120.047659

5. Curtin KM, Pawloski LR, Mitchell P, Dunbar J. COVIDL19 and Morbid Obesity: Associations and Consequences for Policy and Practice. World Medical & Health Policy. 2020 Aug 9. doi: 10.1002/wmh3.361

6. University of Alabama in Tuscaloosa. Relationship between COVID-19 deaths and morbid obesity. ScienceDaily. Retrieved October 17, 2020 from www.sciencedaily.com/releases/2020/08/200820143854.htm

7. Frühbeck G, Toplak H, Woodward E, Yumuk V, Maislos M, Oppert JM. Obesity: the gateway to ill health-an EASO position statement on a rising public health, clinical and scientific challenge in Europe. Obesity facts. 2013;6(2):117–20. doi: 10.1159/000350627

8. Goossens GH, Dicker D, Farpour-Lambert NJ, Frühbeck G, Mullerova D, Woodward E, Holm JC. Obesity and COVID-19: A Perspective from the European Association for the Study of Obesity on Immunological Perturbations, Therapeutic Challenges, and Opportunities in Obesity. Obesity Facts. 2020;13(4):439–52. https://doi.org/10.1159/000510719

9. Van Kerkhove MD, Vandemaele KA, Shinde V, Jaramillo-Gutierrez G, Koukounari A, Donnelly CA, Carlino LO, Owen R, Paterson B, Pelletier L, Vachon J. Risk factors for severe outcomes following 2009 influenza A (H1N1) infection: a global pooled analysis. PLoS Med. 2011 Jul 5;8(7):e1001053.

10. Sun Y, Wang Q, Yang G, Lin C, Zhang Y, Yang P. Weight and prognosis for influenza A (H1N1) pdm09 infection during the pandemic period between 2009 and 2011: a systematic review of observational studies with meta-analysis. Infectious Diseases. 2016 Dec 1;48(11-12):813–22.

11. Kwok S, Adam S, Ho JH, Iqbal Z, Turkington P, Razvi S, Le Roux CW, Soran H, Syed AA. Obesity: A critical risk factor in the COVIDL19 pandemic. Clinical obesity. 2020 Aug 28:e12403. https://doi.org/10.1111/cob.12403

12. Wadman M. Why COVID-19 is more deadly in people with obesity—even if they’re young. AAAS-Science. Health, Corona Virus. 2020 Sep 8. doi:10.1126/science.abe7010. Retrieved from: https://www.sciencemag.org/news/2020/09/why-covid-19-more-deadly-people-obesity-even-if-theyre-young on 19/10/2020 at 00:21

13. Popkin BM, D.S, Green WD, Beck MA, Algaith T, Herbst CH, Alsukait RF, Alluhidan M, Alazemi N, Shekar M. Individuals with obesity and COVIDL19: A global perspective on the epidemiology and biological relationships. Obesity Reviews. 2020 Nov;21(11):e13128. https://doi.org/10.1111/obr.13128

14. Jones-Smith JC, Gordon-Larsen P, Siddiqi A, Popkin BM. Cross-national comparisons of time trends in overweight inequality by socioeconomic status among women using repeated cross-sectional surveys from 37 developing countries, 1989–2007. American journal of epidemiology. 2011 Mar 15;173(6):667–75.

15. Jones-Smith JC, Gordon-Larsen P, Siddiqi A, Popkin BM. Is the burden of overweight shifting to the poor across the globe? Time trends among women in 39 low-and middle-income countries (1991–2008). International journal of obesity. 2012 Aug;36(8):1114–20.

16. Alwarawrah Y, Kiernan K, MacIver NJ. Changes in nutritional status impact immune cell metabolism and function. Frontiers in immunology. 2018 May 16; 9:1055. doi: 10.3389/fimmu.2018.01055. PMID: 29868016; PMCID: PMC5968375

17. Eliakim A, Swindt C, Zaldivar F, Casali P, Cooper DM. Reduced tetanus antibody titers in overweight children. Autoimmunity. 2006 Jan 1;39(2):137–41. doi: 10.1080/08916930600597326

18. Neidich SD, Green WD, Rebeles J, Karlsson EA, Schultz-Cherry S, Noah TL, Chakladar S, Hudgens MG, Weir SS, Beck MA. Increased risk of influenza among vaccinated adults who are obese. International journal of obesity. 2017 Sep;41(9):1324–30.

19. Rahman S, Singh K, Dhingra S, Charan J, Sharma P, Islam S, Jahan D, Iskandar K, Samad N, Haque M. The Double Burden of the COVID-19 Pandemic and Polypharmacy on Geriatric Population–Public Health Implications. Therapeutics and Clinical Risk Management. 2020 Oct 20; 16:1007–22.

20. Docherty AB, Harrison EM, Green CA, Hardwick HE, Pius R, Norman L, Holden KA, Read JM, Dondelinger F, Carson G, Merson L. Features of 16,749 hospitalised UK patients with COVID-19 using the ISARIC WHO Clinical Characterisation Protocol. medRxiv. 2020 Jan 1. doi: 10.1101/2020.04.23.20076042.

21. Caussy C, Wallet F, Laville M, Disse E. Obesity is associated with severe forms of COVID□19. Obesity. 2020 Apr 21; 28: 1175. doi:10.1002/oby.22842

22. Tartof SY, Qian L, Hong V, Wei R, Nadjafi RF, Fischer H, Li Z, Shaw SF, Caparosa SL, Nau CL, Saxena T. Obesity and mortality among patients diagnosed with COVID-19: results from an integrated health care organization. Annals of internal medicine. 2020 Aug 12.

23. Herbst HC, Alsikart FR, Shekar M, El-Saharty S, Menon R. Obesity and COVID-19: A renewed call to address a growing crisis. Investing in health – Worldbank blogs. Retrieved from: https://blogs.worldbank.org/health/obesity-and-covid-19-renewed-call-address-growing-crisis on 18/10/2020 at 11:38am.

24. Braun ES, Crawford FW, Desai MM, Meek J, Kirley PD, Miller L, Anderson EJ, Oni O, Ryan P, Lynfield R, Bargsten M. Obesity not associated with severity among hospitalized adults with seasonal influenza virus infection. Infection. 2015 Oct 1;43(5):569–75. doi: 10.1007/s15010-015-0802-x.

25. Morgan OW, Bramley A, Fowlkes A, Freedman DS, Taylor TH, Gargiullo P, Belay B, Jain S, Cox C, Kamimoto L, Fiore A. Morbid obesity as a risk factor for hospitalization and death due to 2009 pandemic influenza A (H1N1) disease. PloS one. 2010 Mar 15;5(3):e9694.

26. Umbrello M, Fumagalli J, Pesenti A, Chiumello D. Pathophysiology and management of acute respiratory distress syndrome in obese patients. InSeminars in respiratory and critical care medicine 2019 Feb (Vol. 40, No. 01, pp. 040–056). Thieme Medical Publishers.

27. Simonnet A, Chetboun M, Poissy J, Raverdy V, Noulette J, Duhamel A, Labreuche J, Mathieu D, Pattou F, Jourdain M, LICORN and the Lille COVID□19 and Obesity study group. High prevalence of obesity in severe acute respiratory syndrome coronavirus□2 (SARS□CoV□2) requiring invasive mechanical ventilation. Obesity. 2020 Apr 9. https://doi.org/10.1002/oby.22831

28. Kalligeros M, Shehadeh F, Mylona EK, Benitez G, Beckwith CG, Chan PA, Mylonakis E. Association of Obesity with Disease Severity among Patients with COVIDL19. Obesity (Silver Spring, Md.). 2020 Apr 30. doi: 10.1002/oby.22859.

29. Hamer M, Gale CR, Kivimåki M, Batty GD. Overweight, obesity, and risk of hospitalization for COVID-19: A community-based cohort study of adults in the United Kingdom. Proceedings of the National Academy of Sciences. 2020 Sep 1;117(35):21011–3.

30. Seiglie J, Platt J, Cromer SJ, Bunda B, Foulkes AS, Bassett IV, Hsu J, Meigs JB, Leong A, Putman MS, Triant VA. Diabetes as a Risk Factor for Poor Early Outcomes in Patients Hospitalized With COVID-19. Diabetes care. 2020 Aug 25; 28(7):1195–99. doi: 10.1002/oby.22831

31. Palaiodimos L, Kokkinidis DG, Li W, Karamanis D, Ognibene J, Arora S, Southern WN, Mantzoros CS. Severe obesity, increasing age and male sex are independently associated with worse in-hospital outcomes, and higher in-hospital mortality, in a cohort of patients with COVID-19 in the Bronx, New York. Metabolism. 2020 Jul; 108:154262. doi: 10.1016/j.metabol.2020.154262

32. Cai Q, Chen F, Wang T, Luo F, Liu X, Wu Q, He Q, Wang Z, Liu Y, Liu L, Chen J. Obesity and COVID-19 severity in a designated hospital in Shenzhen, China. Diabetes care. 2020 Jul 1;43(7):1392–8. https://doi.org/10.2337/dc20-0576

33. Gao F, Zheng KI, Wang XB, Sun QF, Pan KH, Wang TY, Chen YP, Targher G, Byrne CD, George J, Zheng MH. Obesity is a risk factor for greater COVID-19 severity. Diabetes Care. 2020 May 12; 43(7):E72–4. https://doi.org/10.2337/dc20-0682

34. Nakeshbandi M, Maini R, Daniel P, Rosengarten S, Parmar P, Wilson C, Kim JM, Oommen A, Mecklenburg M, Salvani J, Joseph MA. The impact of obesity on COVID-19 complications: a retrospective cohort study. International Journal of Obesity. 2020 Sep;44(9):1832–7. https://doi.org/10.1038/s41366-020-0648-x

35. Docherty AB, Harrison EM, Green CA, Hardwick HE, Pius R, Norman L, Holden KA, Read JM, Dondelinger F, Carson G, Merson L. Features of 20 133 UK patients in hospital with covid-19 using the ISARIC WHO Clinical Characterisation Protocol: Prospective Observational Cohort Study. BMJ. 2020 May 22;369. https://doi.org/10.1136/bmj.m1985

36. Huang R, Zhu L, Xue L, Liu L, Yan X, Wang J, Zhang B, Xu T, Ji F, Zhao Y, Cheng J. Clinical findings of patients with coronavirus disease 2019 in Jiangsu province, China: A retrospective, multi-center study. PLOS Neglected Tropical Diseases. 2020 May 8;14(5):e0008280. https://doi.org/10.1371/journal.pntd.0008280

37. Kang Z, Luo S, Gui Y, Zhou H, Zhang Z, Tian C, Zhou Q, Wang Q, Hu Y, Fan H, Hu D. Obesity is a potential risk factor contributing to clinical manifestations of COVID-19. International Journal of Obesity. 2020 Sep 13:1–7. https://doi.org/10.1038/s41366-020-00677-2

38. Petrilli CM, Jones SA, Yang J, Rajagopalan H, O’Donnell L, Chernyak Y, Tobin KA, Cerfolio RJ, Francois F, Horwitz LI. Factors associated with hospital admission and critical illness among 5279 people with coronavirus disease 2019 in New York City: prospective cohort study. BMJ. 2020 May 22;369. https://doi.org/10.1136/bmj.m1966

39. Yadav R, Aggarwal S, Singh A. SARS-CoV-2-host dynamics: Increased risk of adverse outcomes of COVID-19 in obesity. Diabetes & Metabolic Syndrome: Clinical Research & Reviews. 2020 Sep 1;14(5):1355–60. https://doi.org/10.1016/j.dsx.2020.07.030

40. Maier HE, Lopez R, Sanchez N, Ng S, Gresh L, Ojeda S, Burger-Calderon R, Kuan G, Harris E, Balmaseda A, Gordon A. Obesity increases the duration of influenza A virus shedding in adults. The Journal of infectious diseases. 2018 Sep 22;218(9):1378–82.

41. Cocoros NM, Lash TL, DeMaria Jr A, Klompas M. Obesity as a risk factor for severe influenza□like illness. Influenza and other respiratory viruses. 2014 Jan;8(1):25–32. https://doi.org/10.1111/irv.12156

42. Fezeu L, Julia C, Henegar A, Bitu J, Hu FB, Grobbee DE, Kengne AP, Hercberg S, Czernichow S. Obesity is associated with higher risk of intensive care unit admission and death in influenza A (H1N1) patients: a systematic review and metaLanalysis. Obesity Reviews. 2011 Aug;12(8):653–9. https://doi.org/10.1111/j.1467-789X.2011.00864.x

43. Louie JK, Acosta M, Samuel MC, Schechter R, Vugia DJ, Harriman K, Matyas BT. A novel risk factor for a novel virus: obesity and 2009 pandemic influenza A (H1N1). Clinical Infectious Diseases. 2011 Feb 1;52(3):301–12.

44. Nguyen-Van-Tam JS, Openshaw PJ, Hashim A, Gadd EM, Lim WS, Semple MG, Read RC, Taylor BL, Brett SJ, McMenamin J, Enstone JE. Risk factors for hospitalisation and poor outcome with pandemic A/H1N1 influenza: United Kingdom first wave (May– September 2009). Thorax. 2010 Jul 1;65(7):645–51. https://doi.org/10.1136/thx.2010.135210

45. Kassir R. Risk of COVID□19 for patients with obesity. Obesity Reviews. 2020 Jun;21(6). https://doi.org/10.1111/obr.13034

