## Supplementary file for "Obesity as a predictor for adverse outcomes among COVID-19 patients: A meta-analysis"

**Supplementary tables**

Table S1: PubMed search results for obesity and adverse outcomes in COVID-19 patients.

| **Search strategy** | **Results** |
| --- | --- |
| "Obesity" AND (COVID-19 or 2019-nCoV or Coronavirus or SARS-CoV-2) AND ("ICU admission" OR "Hospitalization" OR "Disease severity" OR "Invasive mechanical ventilator" OR "Death" OR "Mortality")  **Timespan**: 1/12/2019 to 2/10/2020, **Article type**: Journal article, **Language**: English | 340 |

**Table S2:** Google Scholar search results for obesity and adverse outcomes in COVID-19 patients.

| **Search strategy** | **Results** |
| --- | --- |
| "Obesity" AND (COVID-19 or 2019-nCoV or Coronavirus or SARS-CoV-2) AND ("ICU admission" OR "Hospitalization" OR "Disease severity" OR "Invasive mechanical ventilator" OR "Death" OR "Mortality")  **Timespan**: 2019 to 2020 | 2600 |

**Table S3:** ScienceDirect search results for obesity and adverse outcomes in COVID-19 patients.

| **Search Strategy** | **Results** |
| --- | --- |
| "Obesity" AND (COVID-19 or 2019-nCoV or Coronavirus or SARS-CoV-2) AND ("ICU admission" OR "Hospitalization" OR "Disease severity" OR "Invasive mechanical ventilator" OR "Death" OR "Mortality")  **Article type:** Research article | 58 |

**Table S4.** Newcastle-Ottawa scale assessment of study quality for **cohort study.**

| Author | Selection | | | | Comparability | | Outcome | | | Study Quality |
| --- | --- | --- | --- | --- | --- | --- | --- | --- | --- | --- |
|  | 1 | 2 | 3 | 4 | 5A | 5B | 6 | 7 | 8 |  |
|  | Exposed cohort truly representative | Non-exposed cohort drawn from the same community | Ascertainment of exposure | Outcome of interest not present at start | Cohorts comparable on basis of age | Cohorts comparable on other factor(s) | Quality of outcome assessment | Follow-up long enough for outcomes to occur | Complete accounting for cohorts |  |
| Simonnet *et al.*, 2020 | * | * | * | * | * |  | * | * | * | 7 |
| Kalligeros *et al.*, 2020 | * | * | * | * | * | * | * | * | * | 9 |
| Hamer *et al.*, 2020 | * | * | * | * |  |  | * | * | * | 7 |
| Seiglie *et al.*, 2020 | * | * | * | * |  |  | * | * | * | 7 |
| Palaiodimos *et al.*, 2020 | * | * | * | * |  |  | * | * | * | 7 |

**Supplementary figures**

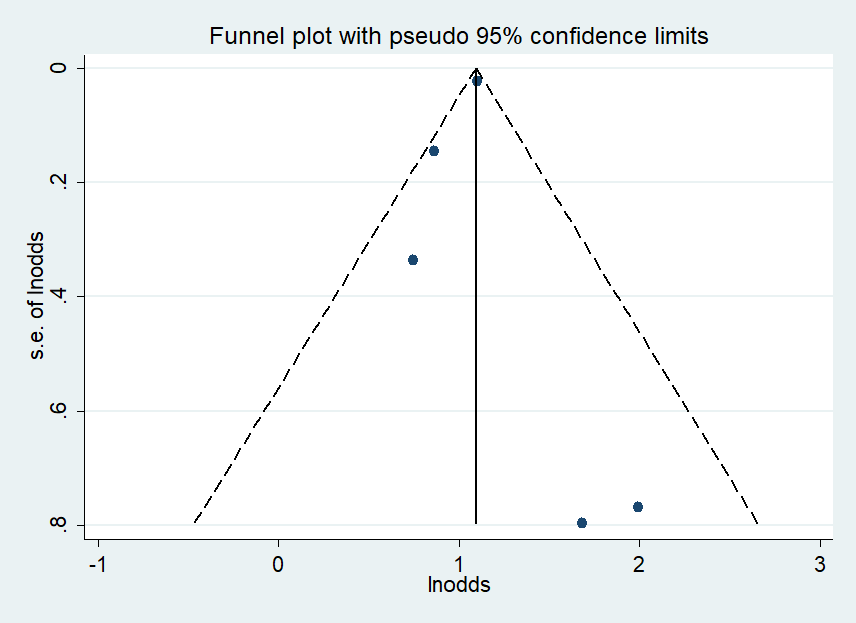

Figure S1: Funnel plot for obesity and adverse outcome on COVID-19 infected persons.

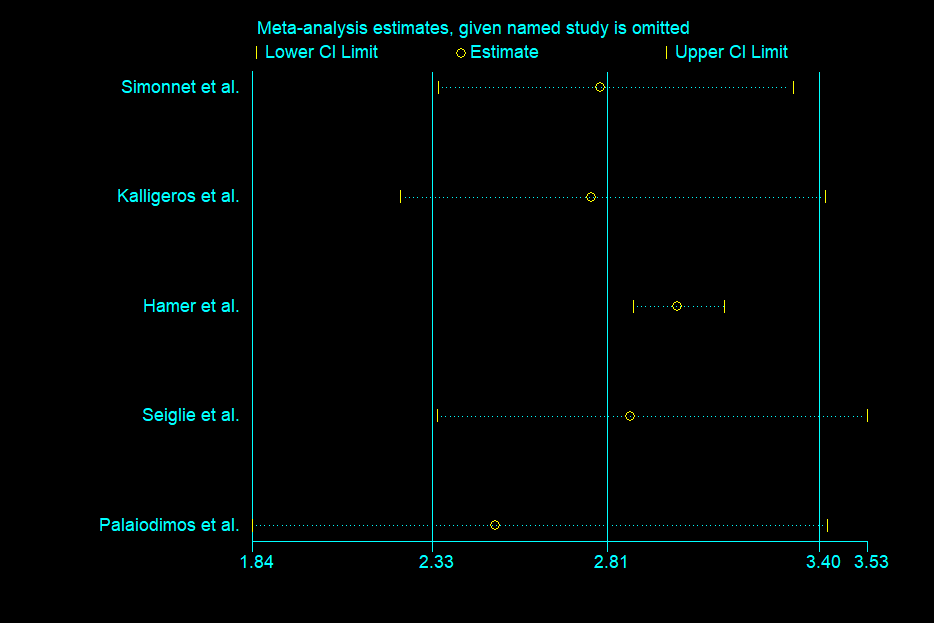

Figure S2: Sensitivity analysis of the involved studies
